# Management of Risk Factors for Health Care Workers After Initial Contact with Patients Infected with SARS-COV-2 in Niamey, The Center of the Epidemic in Niger

**DOI:** 10.1101/2023.03.01.23286420

**Authors:** Hamidou Lazoumar Ramatoulaye, Aliou Sanda Abdal-Kader, Adamou Lagare, Mahamadou A Maiga, Fakani Aboutalib Aliane, François Comlan Aida Sylviane, Idi Issa, Bibata Abdou Sidikou, Garda Idé Oumarou, Ali sidiki, Zeinabou Abdou Aouta, Amina Moussa, Zeinabou Dioffo Alassan, Ibrahim Karidjo, Goni Alhassan Maman Bachir, Issifou Djibo, Salia Moussa, Ibrahim Maman Laminou, Ronan Jambou

**Author notes:** Corresponding author : HAMIDOU LAZOUMAR Ramatoulaye (H.L.R).

## Abstract

**Background:** The covid-19 pandemic is caused by a new corona virus called SARS Cov-2. Health care workers are particularly exposed target.

**Aim:** this study aims to analyze the risk factors of SARS-Cov-2 infection in health workers who have been in contact with positive patient.

**Methods:** This is a prospective cohort conducted among health workers from March 2022 to January 2021 in health care facilities in Niamey. A questionnaire was administrated at inclusion. rRT-PCR was performed if clinical signs were present. ELISA testing was performed at baseline, day15 and day 30. The chi-square or Fisher test, Kaplan-Meir survival model, Cow regression and logistic regression were used as statistical test.

**Results:** 259 health workers were included. More than half of the respondents were female. 45.95% of the participants were nurses and 36.68% were physicians. The prevalence of Covid-19 was 28.8%. 56.4% of the participants had positive serology at day 30. The risk factors associated with Covid-19 were professional category (p=0.024). Membership structure (p<0.001) and the chronic liver disease (p=0.034). Hand hygiene (p=0.019) and alcohol-based hand cleaning (p<0.001) protects against the occurrence of SARS-Cov-2 infection. According to the characterization of the preventive measures those who practiced them rarely were associated with a positive rRT-PCR and those who practice them occasionally or most of the time were associated with negative serology and rRT-PCR.

**Conclusion:** Comorbidities, function and affiliation are the main risk factors for Covid-19 and hand hygiene is a protective factor.

## 1. Introduction

The coronavirus (COVID-19) outbreak first detected in the city of Wuhan, China, in December 2019 (1), has causes a global health crisis, despite the implementation of large-scale control measures. It rapidly spread across the different continents of the world. On January 30, 2020, the World Health Organization announced the outbreak of the new coronavirus (Severe Acute Respiratory Syndrome coronavirus 2 - SARS-CoV-2) as a public health emergency of international concern, declaring it a pandemic on March 11, 2020 (2). Since the beginning of this pandemic, the world has seen an increase in cases with a total of 527,971,809 cases and 6,284,871 deaths (3). The African continent is the last to be affected after Europe and United States. Africa recorded its first case officially on February 15, 2020, two months after it was first identified in China (4). The first confirmed case of Covid-19 in Africa was reported in Egypt on February 14 and the second one a day later in Algeria. By March, COVID-19 cases were reported from most of the continent. By April, nearly every country in Africa had reported COVID-19, with hundreds, if not thousands, of cases reported in the hardest-hit countries (5). Niger has been facing the COVID-19 pandemic since Mars 19, 202 when the first case was reported/ All eight regions of the country are affected by the pandemic (6). According to the situation report of who, the virus was first reports in Niamey, before spreading to other parts of Niger. (7). Faced with this COVID-19 pandemic, professionals working in health facilities are particularly exposed and vulnerable to contamination. They are on the front line of the epidemic and experiment a higher risk of contamination and subsequently of transmission to new patients, colleagues and family members (8). In Africa, they face several challenges in the context of coronavirus disease, such as lack of personal protective equipment (PPE), infection, quarantine, discrimination and attacks on them in society, in the same time as a large responsibility in taking care for their relatives and families (9). In some areas, health care workers have accounted for up to 11% of all confirmed COVID-19 cases, with an increasing number of work-related deaths (10). Health workers in health care facilities in Niger have taken a heavy toll with at least 184 declared cases out of 1059 confirmed cases. This could be explained by the lack of effective implementation of standard infection prevention and control (IPC) precaution (6). Moreover, most of the population including health care workers didn’t believe in the reality of the epidemic, when most of the diagnostic facilities were grouped in CERMES. Very few people attended thus dispensaries and CERMES for biological diagnostic.

In the countries with limited resources like Niger, it is thus crucial to determine which practices are the most effective to prevent contamination of health care workers, to subsequently focus on their dissemination.

In Niger, neither the spread of the first cases of SARS-CoV-2 infection in health care settings nor the analysis of risk factors associated with this infection specific to health care workers is clearly documented. This study was initiated to analyze the diffusion of the infection and to determine the risk factors for contracting SARS-CoV-2 by healthcare workers. It focused on healthcare departments in charge of COVID-19 cases management.

## 2. Methods

### 2.1. Study design

A prospective cohort study was conducted from March 2020 to January 2021 among people working in health facilities of Niamey where first cases of COVID-19 were reported.

### 2.2. Site of study

Data were collected in eleven (11) health facilities: the National Hospital of Niamey (HNN), the General Reference Hospital (HGR), the Regional Hospital Poudriére (HRP), the Issaka Gazobi Maternity Hospital (MIG), the Ambulatory Treatment Center (CTA), the Wangari Clinic and the Arahama Cabinet.

### 2.3. Study population and follow-up procedure of workers and relatives

In this study, all health care workers in charge of SARS-CoV-2 infected patients were eligible, whatever their positions, including those who handled blood samples or in contact with biological fluids through cleaning of the surfaces or equipment.

This was an exhaustive sampling including all workers who had contact with a COVID-19 cases and who agreed to participate. Both private and public structures were considered as soon as they were in charge of COVID-19 cases. Date of contact with the patients should be less than one week at the day of inclusion of the worker.

The health workers included in the study were followed for one month during which they were called every day to inquire about their health status (presence or absence of clinical signs in favor of SARS-CoV-2 disease, temperature measurement). All people presenting clinical signs during the month of follow-up were tested by PCR for Sars-Cov-2 detection.

### 2.4. Data and samples collected

At inclusion, a standard questionnaire was completed including information on sociodemographic characteristics, exposure to the SARS-CoV-2 infected patient, personal and collective infection prevention facilities, and laboratory tests used.

Blood samples of 2-3 ml were taken at day 0 (inclusion in the study), day 14 and day 30. As the study was performed at the beginning of the epidemic in Niger and before any vaccination, for risk factor analysis, only serologies at Day 30 were considered to get a complete picture of what happened during the month. Indeed, during follow-up, patients may have experienced asymptomatic infection and/ or should have not report clinic signs for personal reasons and therefore should have not been rRT-PCR tested.

RT-PCR and WANTAI SARS-CoV-2 Ab ELISA (total antibodies Wantai Biological Pharmacy Enterprise o, Ltd, Beijing, China) were respectively used for the diagnosis of COVID-19 and seroprevalence of SARS-CoV-2 antibodies.

The laboratory staff in charge of these different samples had been trained in the safe handling of samples and the implementation of virological confirmation of suspected cases of SARS-CoV-2.

### 2.5. Statistical analysis

Data were collected on Redcap and analyzed with R software (© R-4.2.1). The variables serological status, rRT-PCR result and occurrence of any of the Covid symptoms were defined as dependent variables of the study.

A general descriptive analysis of risk factors for COVID-19 among health workers was first performed and a Chi-square or Fisher test was performed to compare proportions considering a 5% margin of error.

A factorial correspondence analysis was performed with the explanatory variables and then a projection of the dependent variables (serological status and rRT-PCR status) was made on the graphs to visualize the profiles of the health workers at risk.

A Kaplan-Meir survival model was used to determine the probability of COVID-19 manifestation in health workers forty-five (45) days after contact and a Cox regression Cox regression to determine the determining risk factors. A first crude cox model (Model I) without adjustment was performed to determine the risk associated with the explanatory variables, then a second model (Model II) with adjustment on the explanatory variables, and then a final model (model III) by top-down selection, with the threshold determined by the AIC (Akaike information criterion). The conditions of realization of the Cox test were carried out and a stratification was made for the variables not respecting the conditions of proportionality of risk in time.

Logistic regression was performed with the same procedure (model I II and III) to determine the risk factors in the form of odds ratios (OR) with their 95% confidence intervals (CI) associated with COVID-19 serological status.

Because of the failure to perform rRT-PCR in several participants with COVID symptoms, multivariate regression analysis was not performed on this variable.

### 2.6. Ethical consideration

Ethical clearance was obtained from the National Ethics Committee for Health Research (CNERS) with the reference N°04/2020/CNERS during its session on March 12, 2020. Written informed consent was obtained by each participant. Respondents were informed that they had the right to refuse or discontinue their participation in the study at any time. The information collected was kept confidential. To this end, a code was assigned to each health facility, as well as an identification number for all study participants.

## 3. Results

### 3.1. Characteristics of workers included in the study

A total of 288 health workers following inclusion criteria, were registered and 259 were selected for univariate analysis after cleaning of the database. As presented in Table (I), the socio-demographic status revealed that the average age of the participants was 35 years with extremes ranging from 21 years to 65 years. A significant number of workers were women (53.7%). According to the status, senior health technicians were the most represented (46%) followed by physicians (36.7%). Trainees were the least represented in this study (3.5%). Asthma (7.3%), obesity (5%), heart disease (3.8%) and chronic lung disease (2.3%) were the most recorded risk factors .

**Table 1 :**
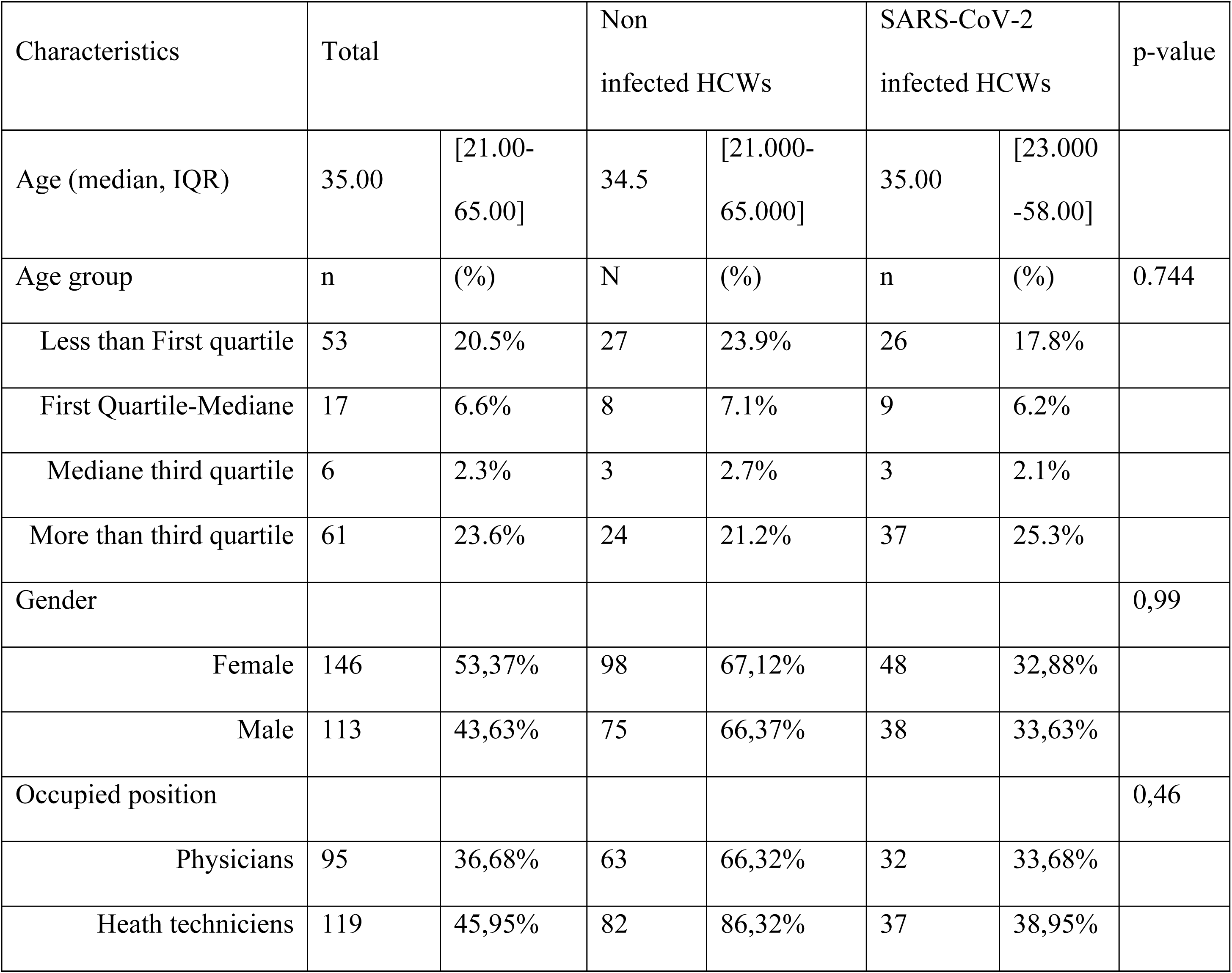
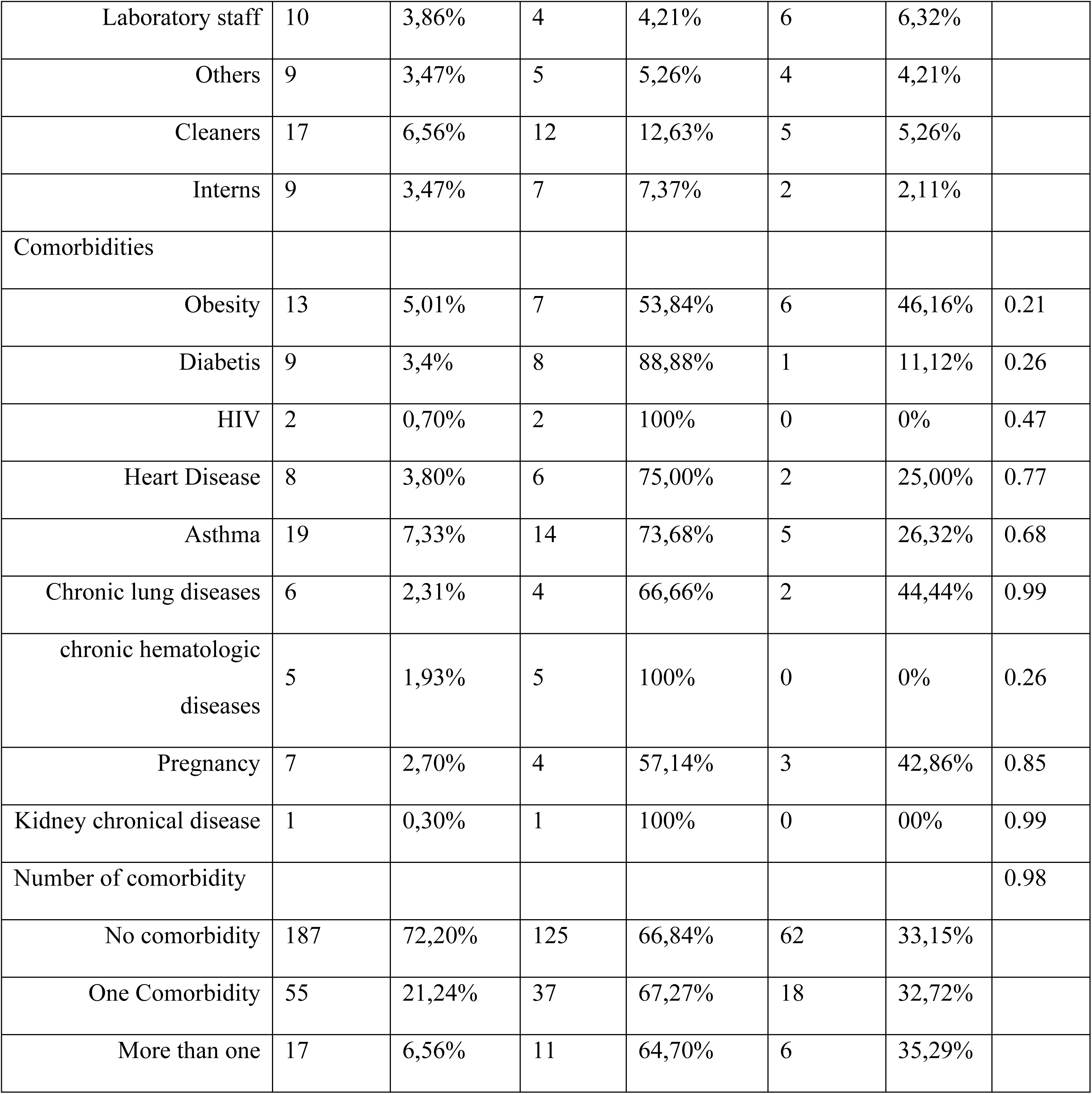
Socio-demographic characterization of the population

### 3.2. Serological follow-up

During the course of the study, the patients were subjected to blood samples on day 0 (baseline), on day 14 and on day 30. For the risk analysis, only serology on day 30 were considered in order to obtain a complete picture on the whole evolution during the first month. Indeed, during follow-up, patients may have experienced asymptomatic infection and/or did not report clinical signs for personal reasons and therefore did not benefit from rRT-PCR analysis. The data presented after day 0 only include workers who were recruited within 15 days after a contact with a COVID-19 positive patient. 170 HCWs were selected for this analysis, out of whom 59 were positive at inclusion. For the 110 negative individuals, 101 were sampled at day14 (with 29 new positive serologies). For the 72 negative at day 14, 74 (including 2 workers not sampled at day 14) were sampled at day 30 with 8 new negatives. The conversion rate decreased from 35 to 28.7 and 10% at day 0, day14 and day 30 respectively. Interestingly, there was no clear difference in the number of symptomatic or CRO+ individuals at each stage between the positive and negative groups. At the end of the follow-up, 50% of the subjects changed their serostatus, out of which only 11% reported clinical signs.

### 3.3. Symptomatic disease and rRT-PCR

One hundred and eighteen (44.40%) of the workers were symptomatic during this one month follow-up. All were rRT-PCR COVID19 tested among whom 34 (28.8%) were positive. Among the positive one, 11 (64.7%) were female and 12 (35.3%) were male. According to the status, medical doctors (17.4%) were the most impacted followed by senior technicians (10.9%).

### 3.4. Risk factors for clinical disease and/or positive rRT-PCR or serology

Risk factor analyses were performed only for participants who were included within 15 days after a contact with SARS-CoV2 positive patients. (Table II) summarizes the results of rRT-PCR or serology positive prevalence and associated factors. 28.8% and 56.47% of the participants were positive for rRT-PCR and serology respectively.

**Table II :**
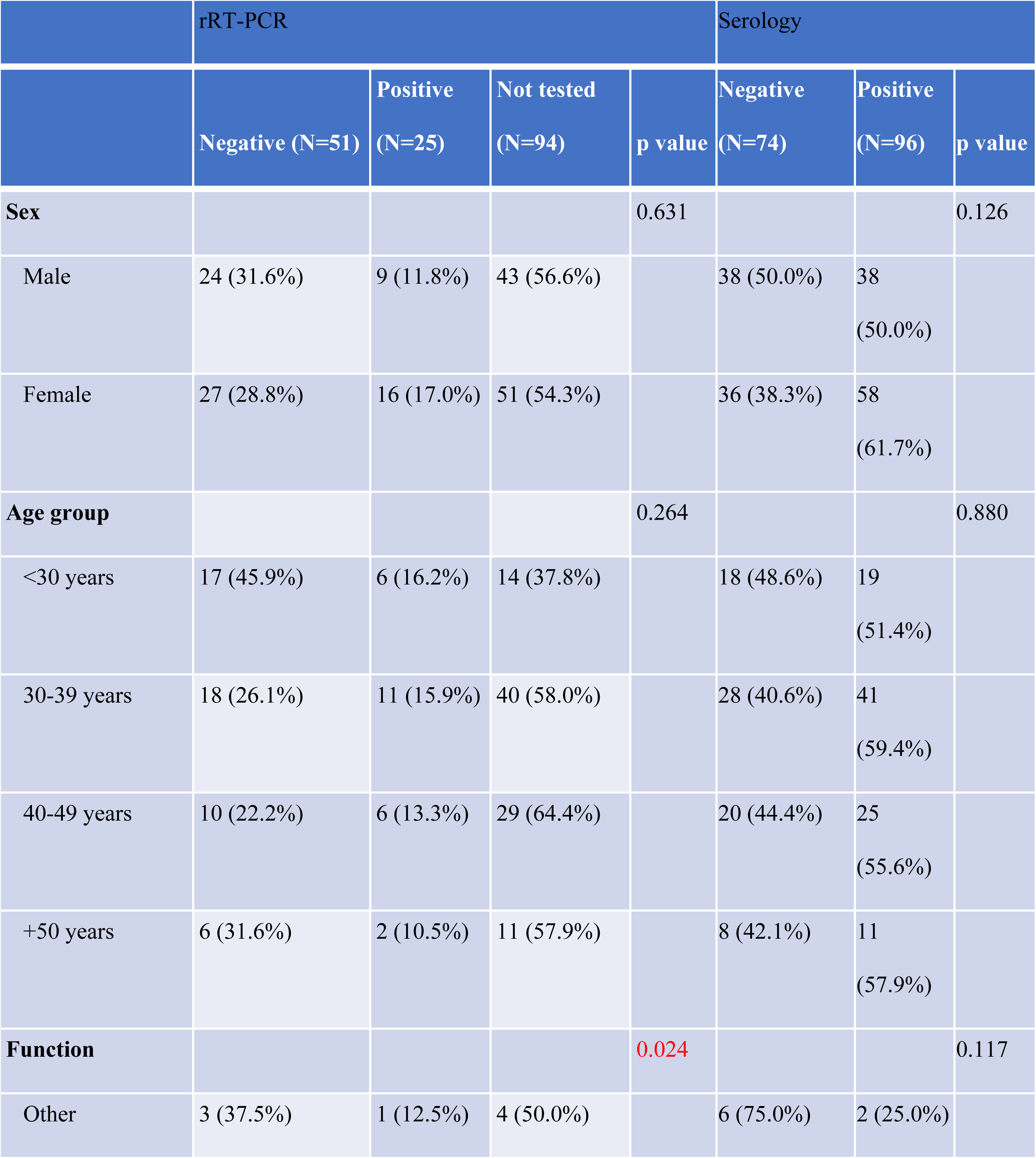
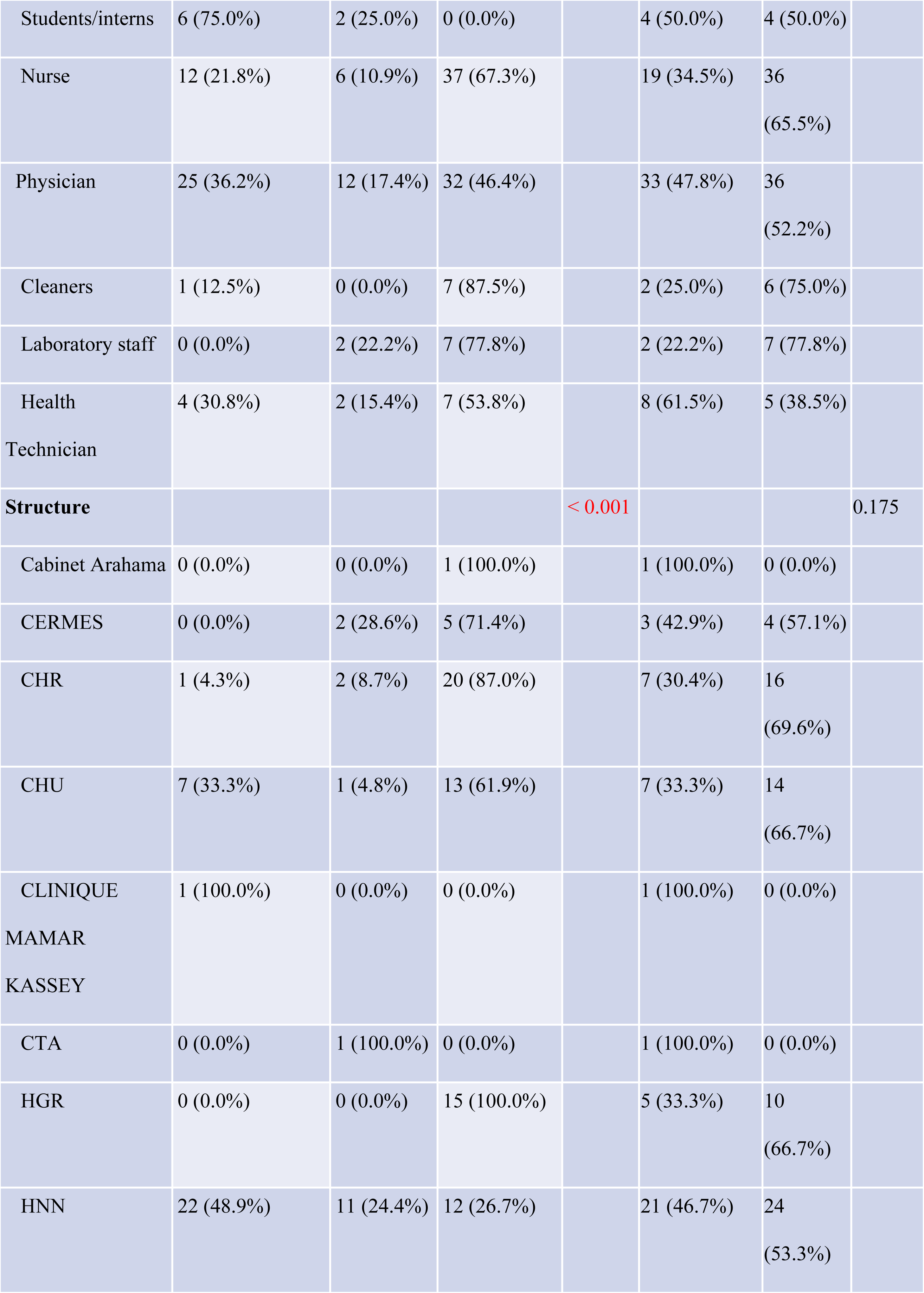
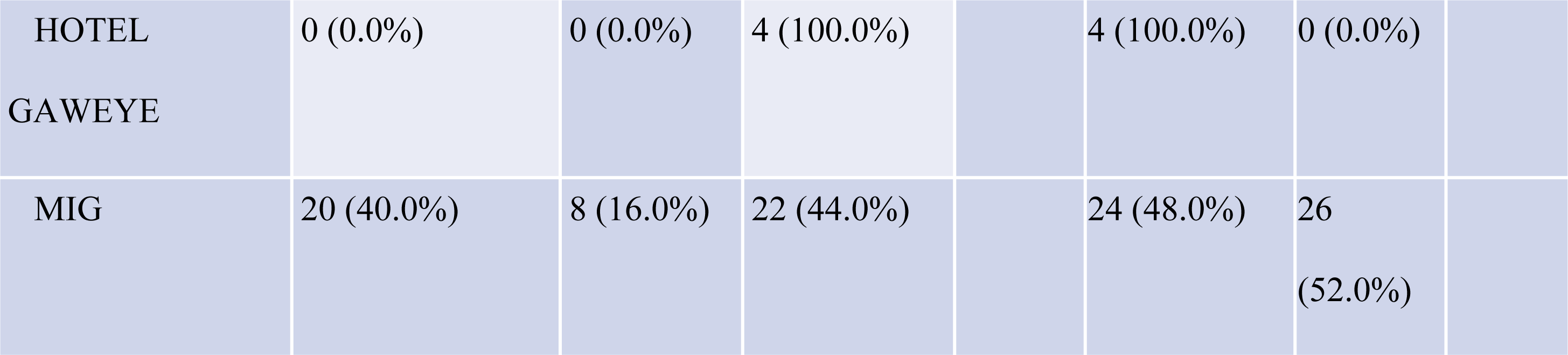
Socio-demographic characteristics of participants at inclusion according-rRT-PCR and serology results.

In univariate analysis (Table II), there was a statistically significant more or less in the proportion of positive rRT-PCR according to the status (p=0.024), the type of health structure (p<0.001).

In the analysis between infection prevention and control practices and rRT-PCR (Table III), the presence of chronic liver disease (p=0.034), the hand washing practice (p=0.019), and hand hygiene using alcohol (p<0.001).

**Table III :**
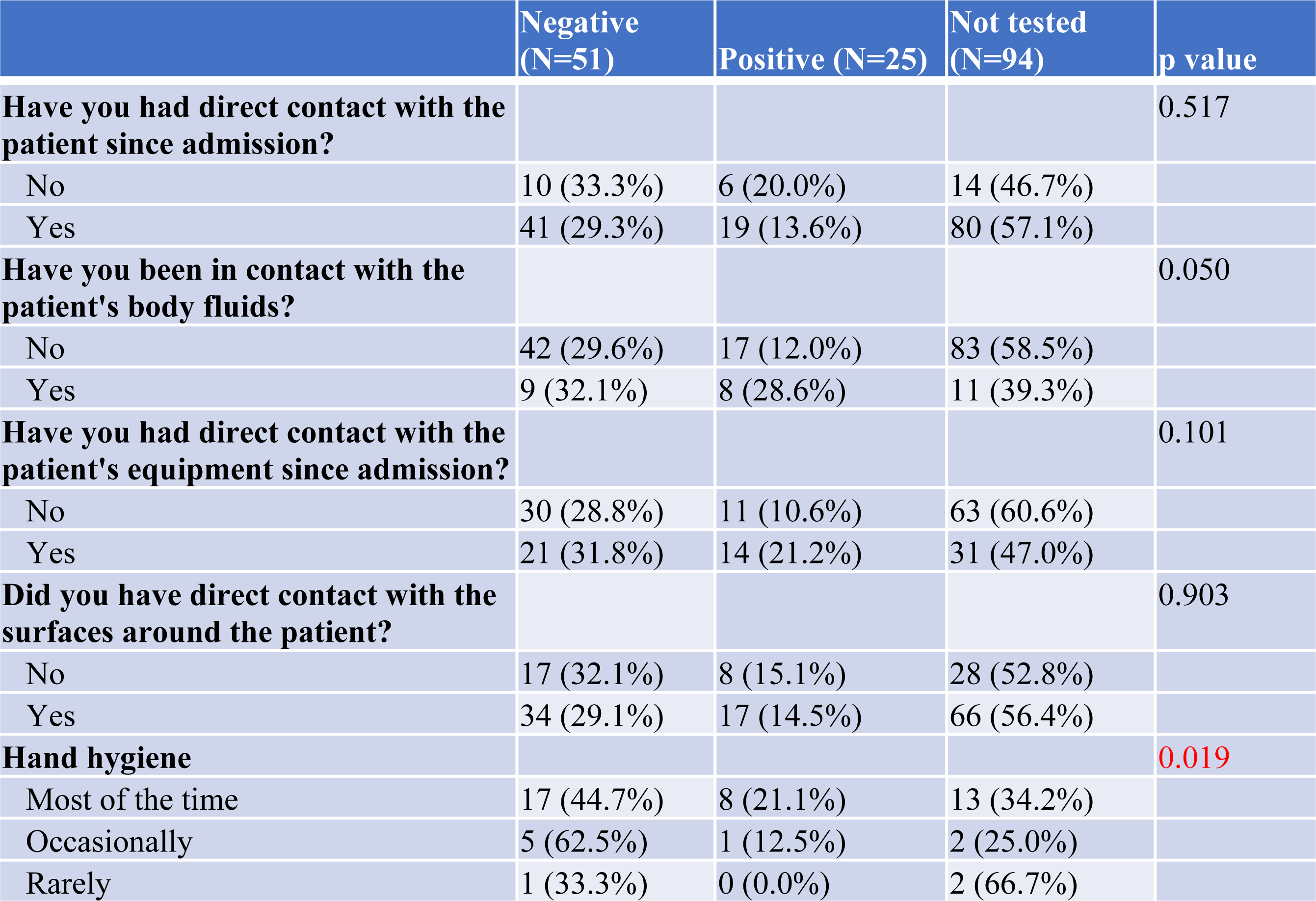
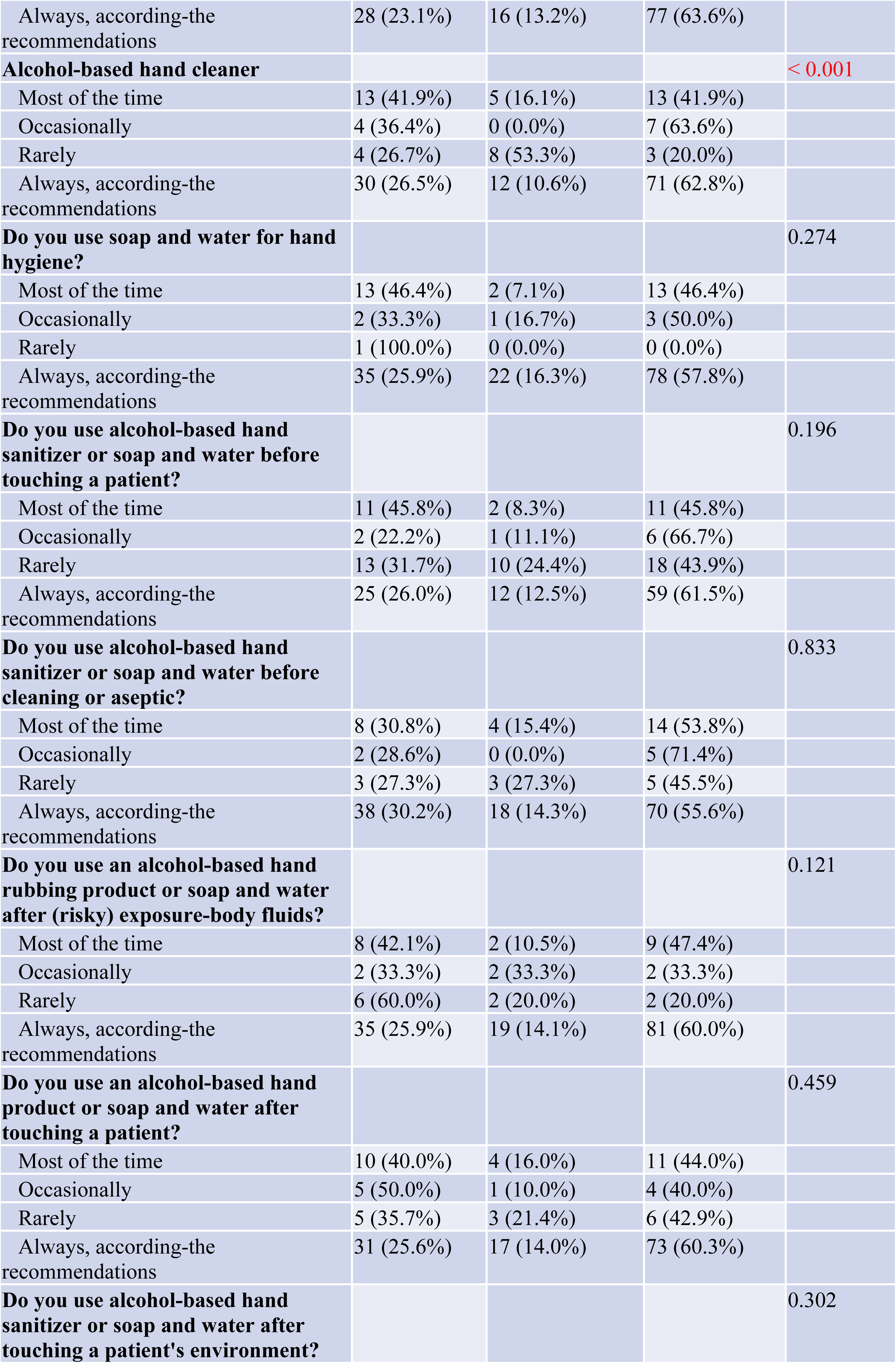
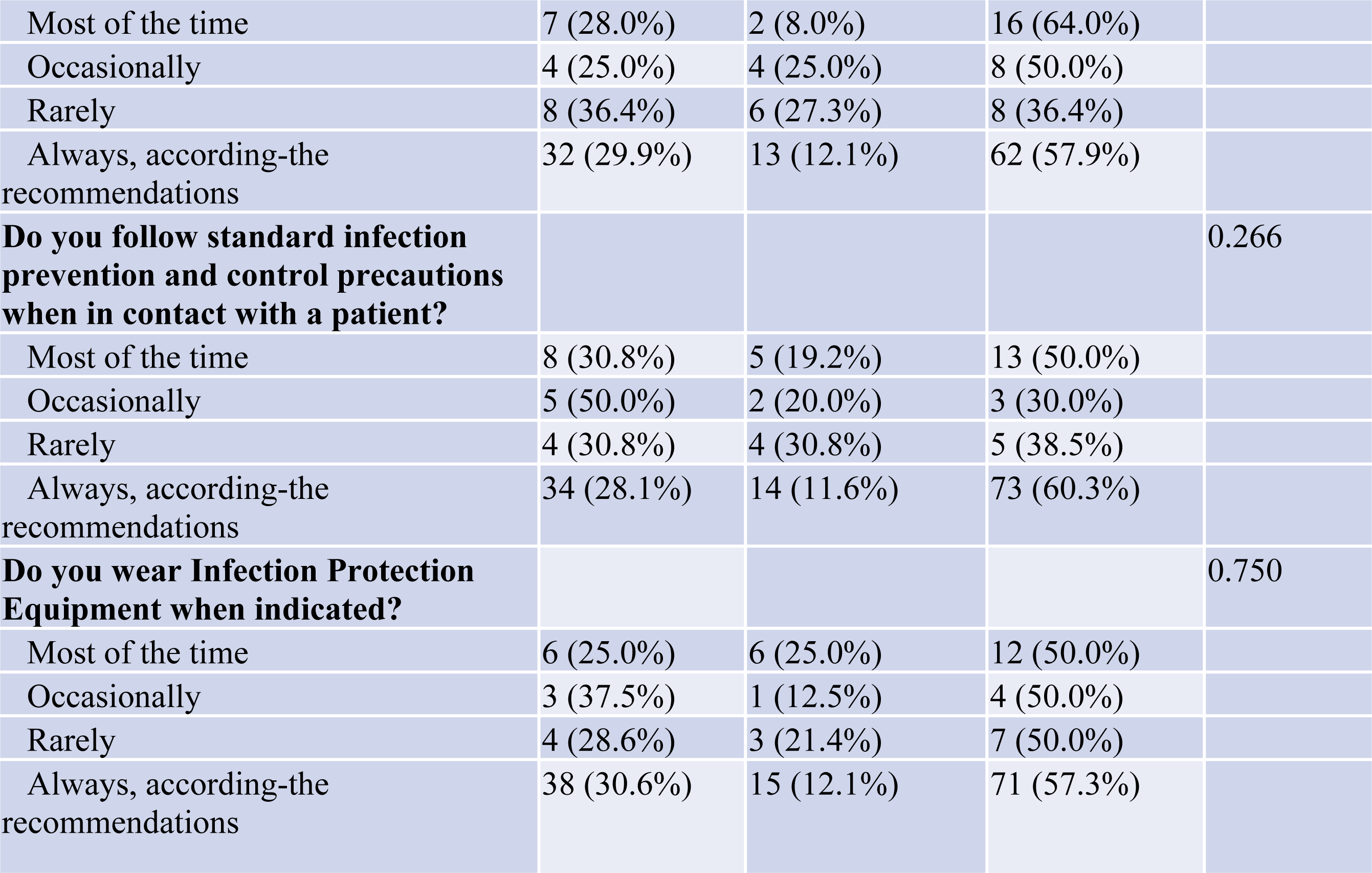
Infection Prevention and Control Practices among Health Workers and rRT-PCR Outcome in Niger

The univariate analysis between covid-19 prevention and control practices among health workers and serology (Table IV), shows that the contact with contaminated materials and the use of protective equipment were almost positively and negatively associated with positive serology respectively (p= 0.069, p=0.068).

**Table IV :**
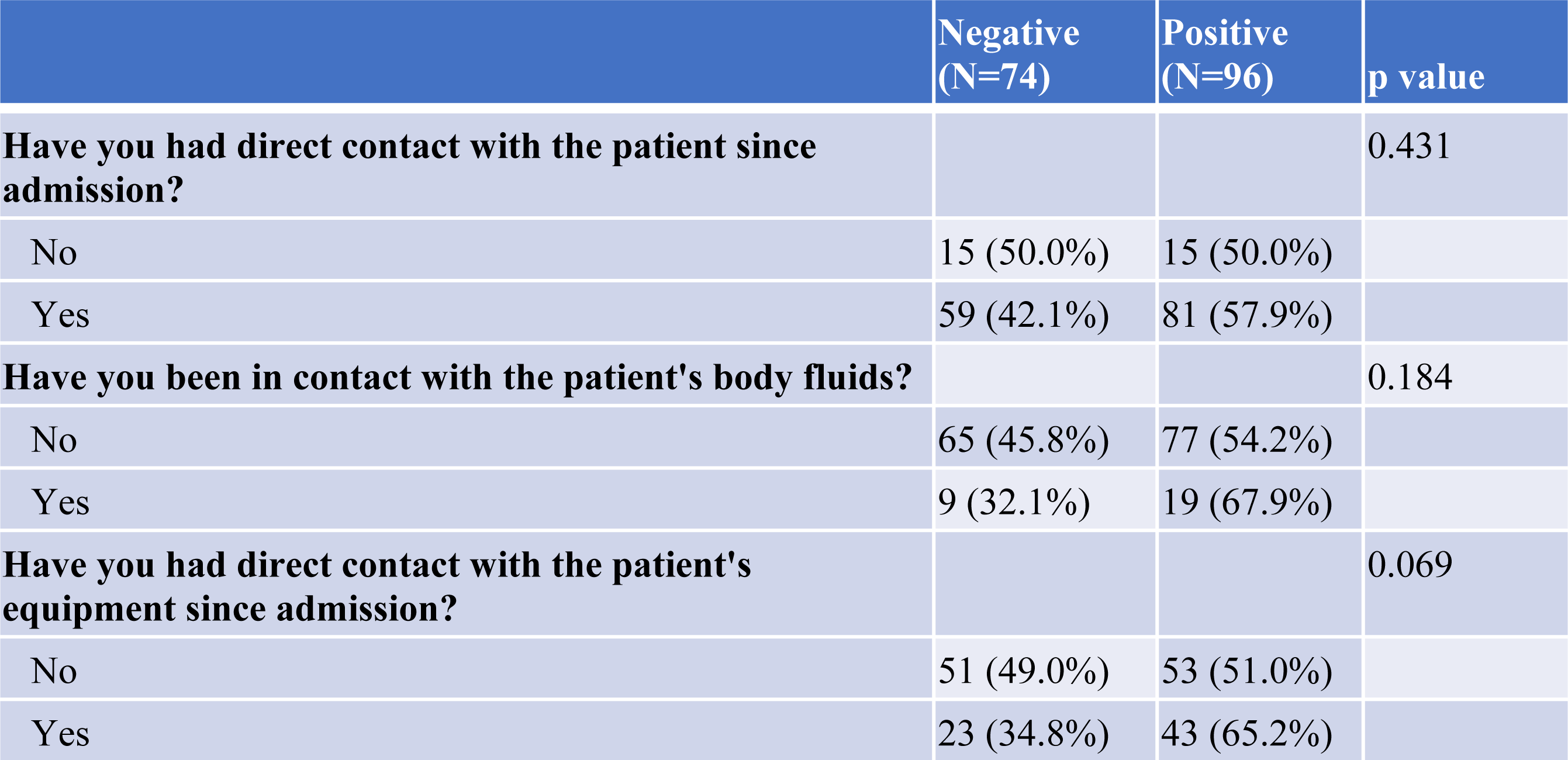
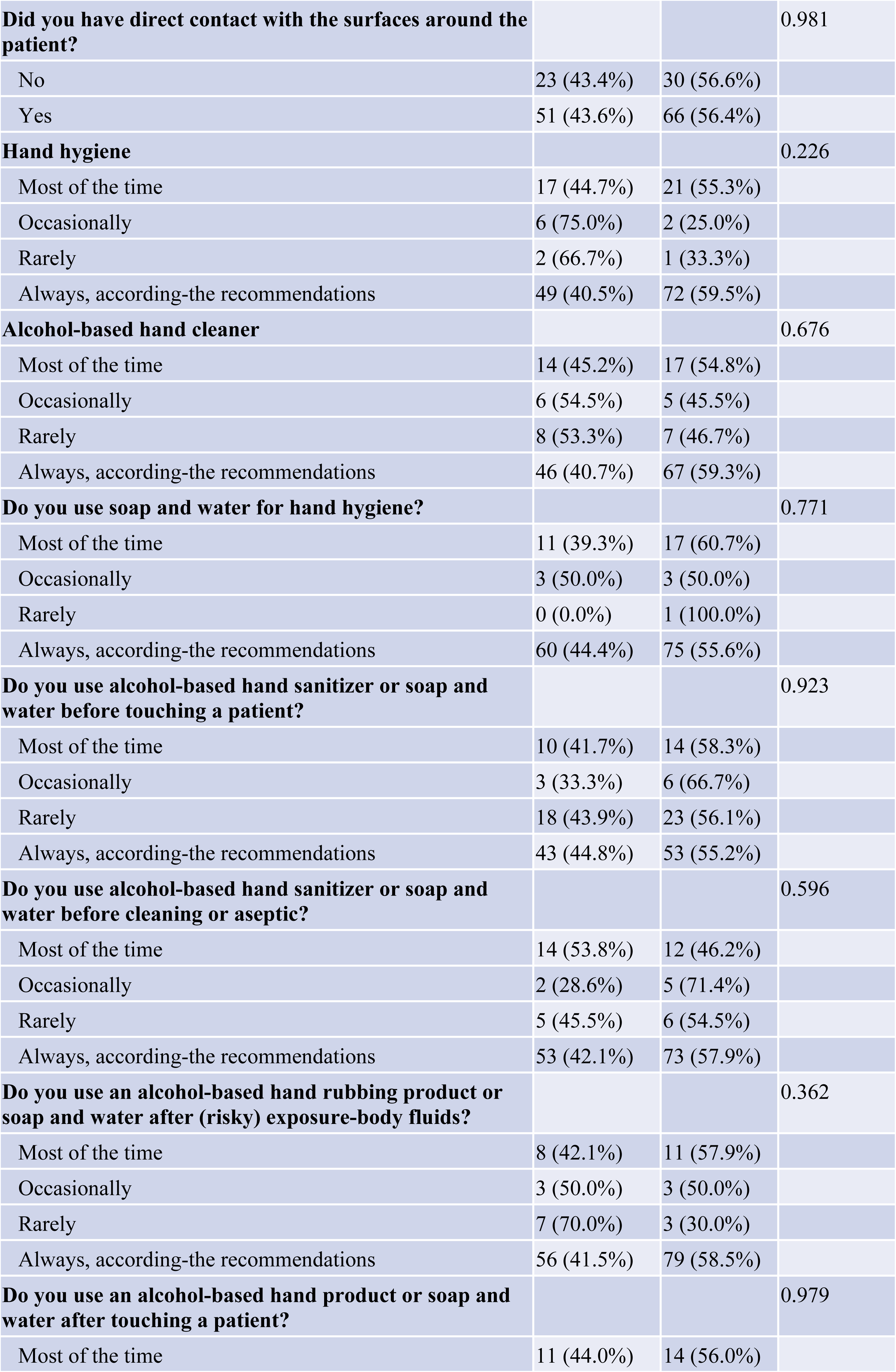
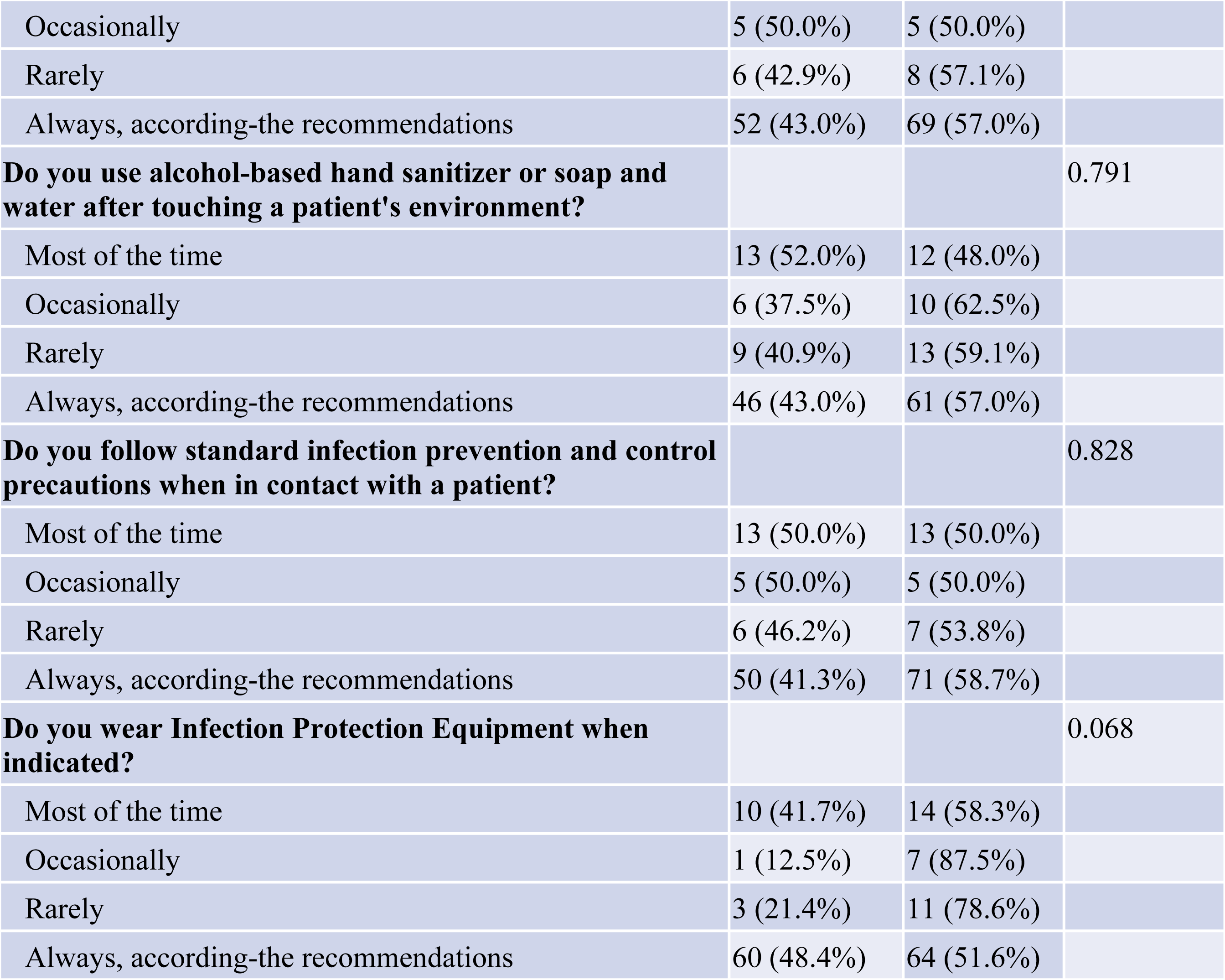
Infection prevention and control practices by health workers and outcome of serology

In the correspondence analysis was performed on dichotomic variables (Fig. 1, 2, 3), the age group < 30 years was the most associated with negative serology. Female, age group 30-39 years, physician occupation (Fig. 3), contact with fluids, contact with medical equipment were the factors the most associated with positive PCR. In the same line, low hygiene practices were associated with positive rRT-PCR, whereas fair hygiene practices were associated with negative rRT-PCR and serology.

**Figure 1 :**
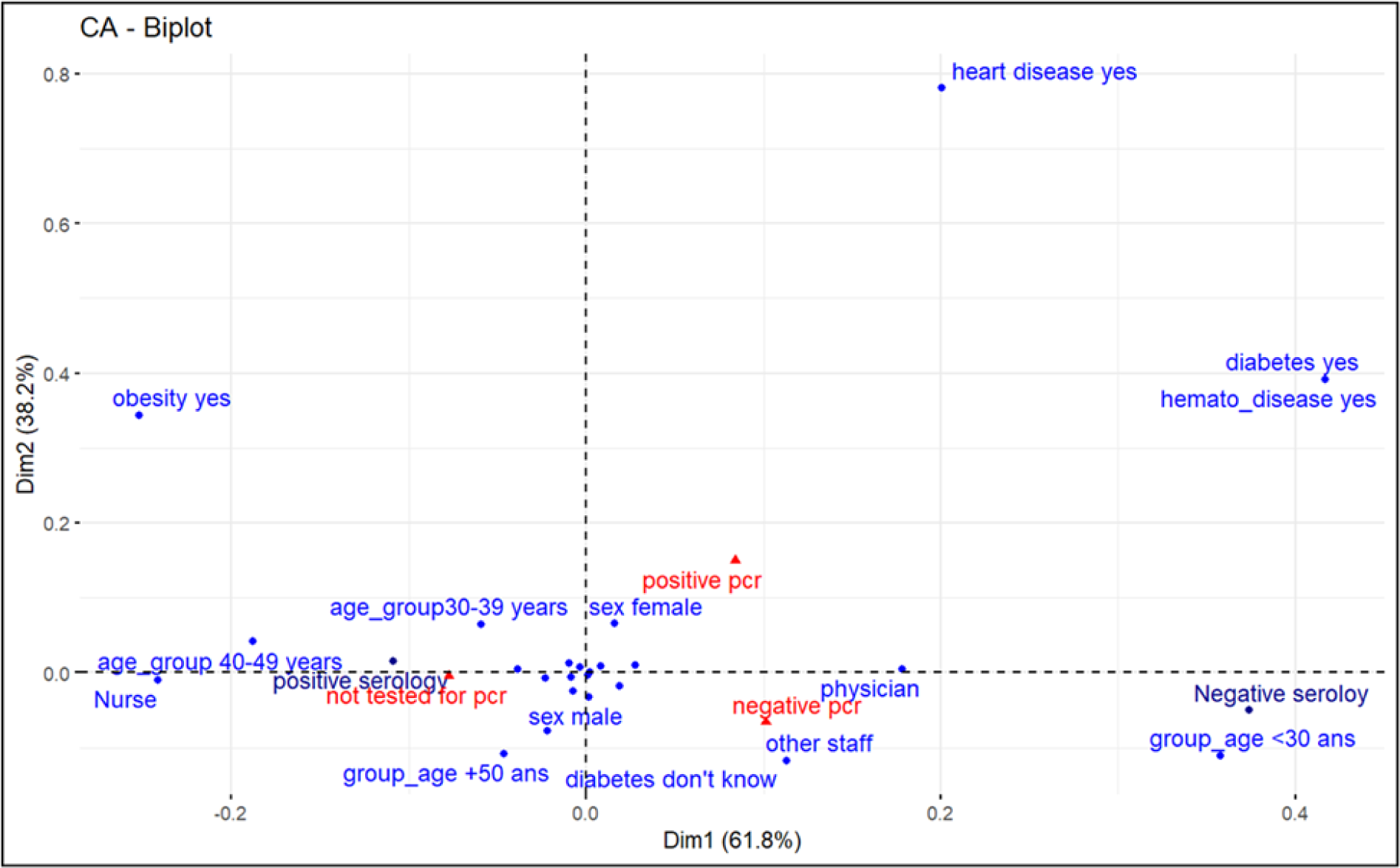
Correspondence analysis of age, function and medical antecedent associated with PCR status and serology

**Figure 2 :**
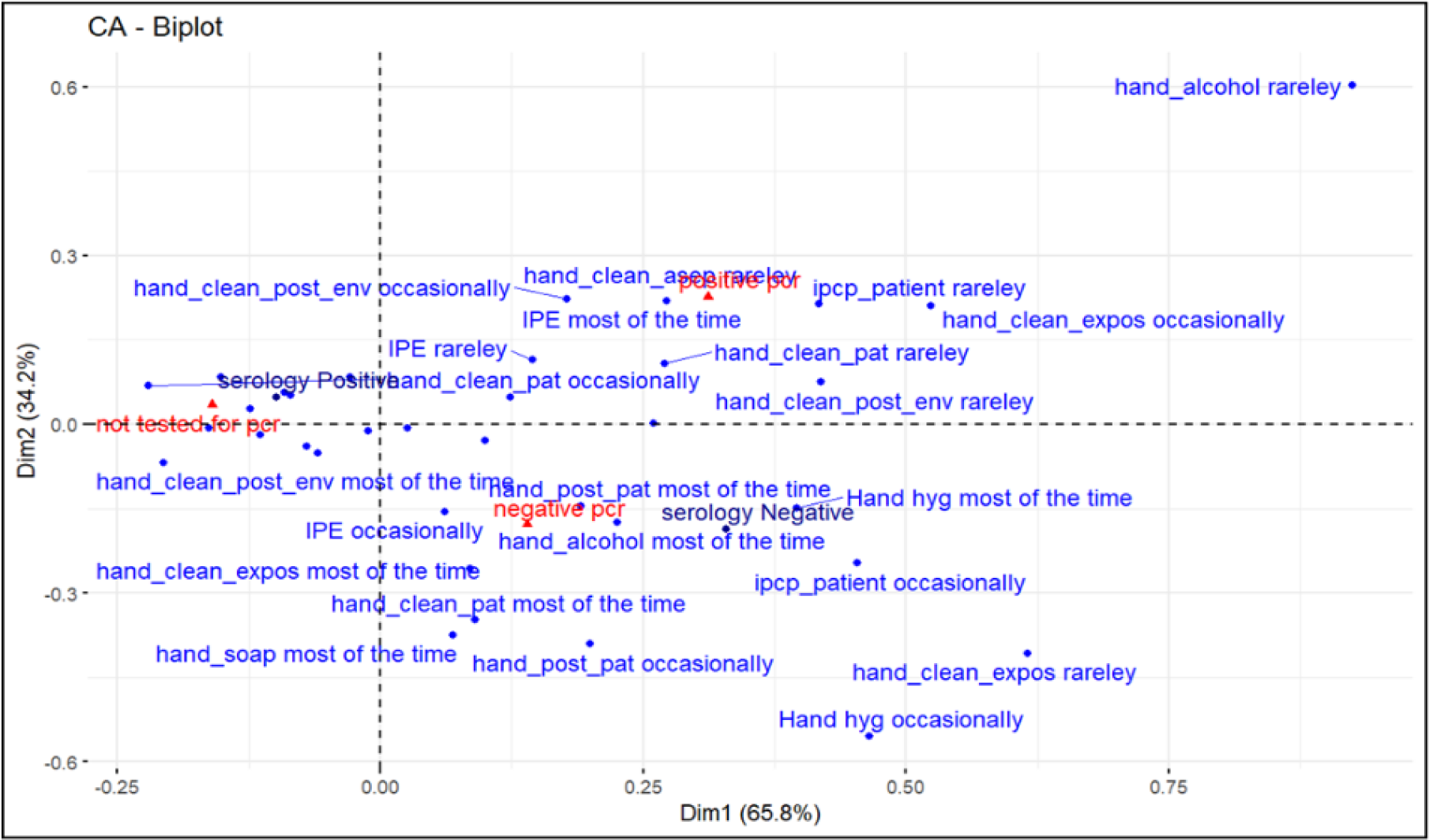
Correspondence analysis of hygiene practices associated with status rRT-PCR and serology

**Figure 3 :**
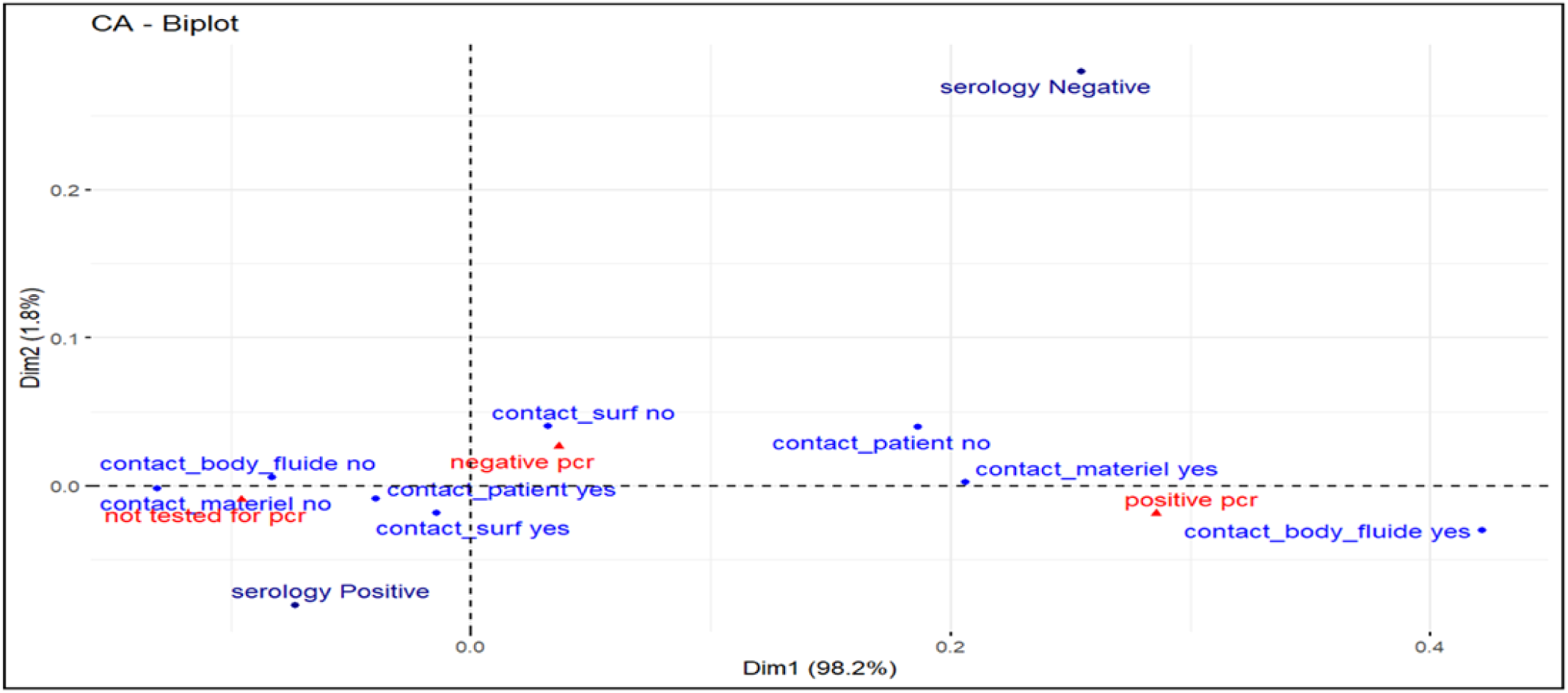
Correspondence analysis of type of exposure associated with rRT-PCR status, serology

The logistic regression analysis focused on risk factors associated with positive serology (Table V). In model I (crude), no factors significantly associated with positive serology. In the model adjusted II, contact with patient’s body fluid was associated with positive serology (OR 95% 5.71 [1.56-24.9]). Participants who don’t know their status about diabetes were less likely to have positive serology compared to the others (OR95% 0.04, [0.00-0.61]). In the final model (III), female gender was a major risk factor for positive serology (OR95% 2.22 [1.09-4.68]) and the use of protective equipment was a protective one (OR 95% 2.88, [0.97-9.36]).

**Table V :**
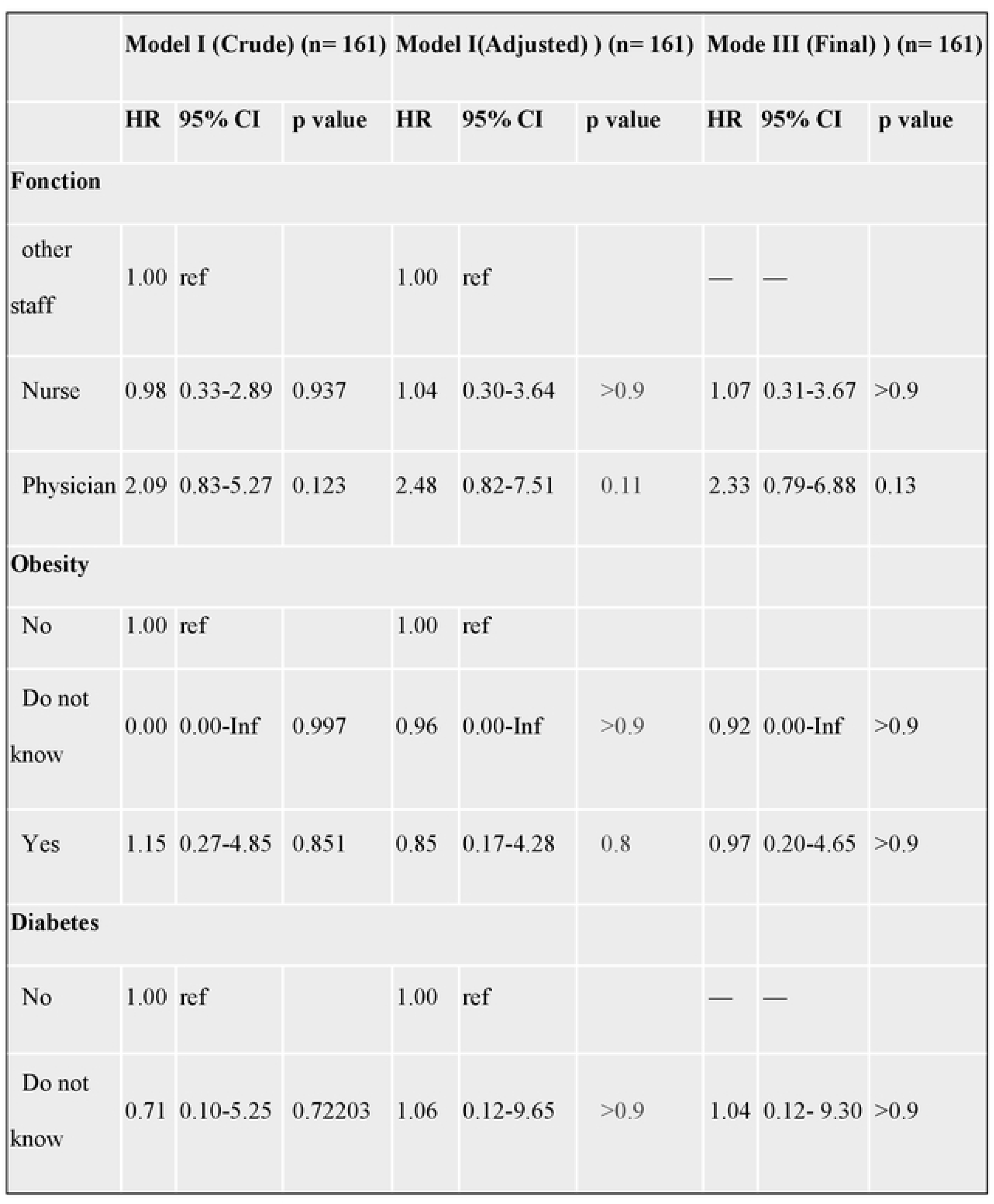
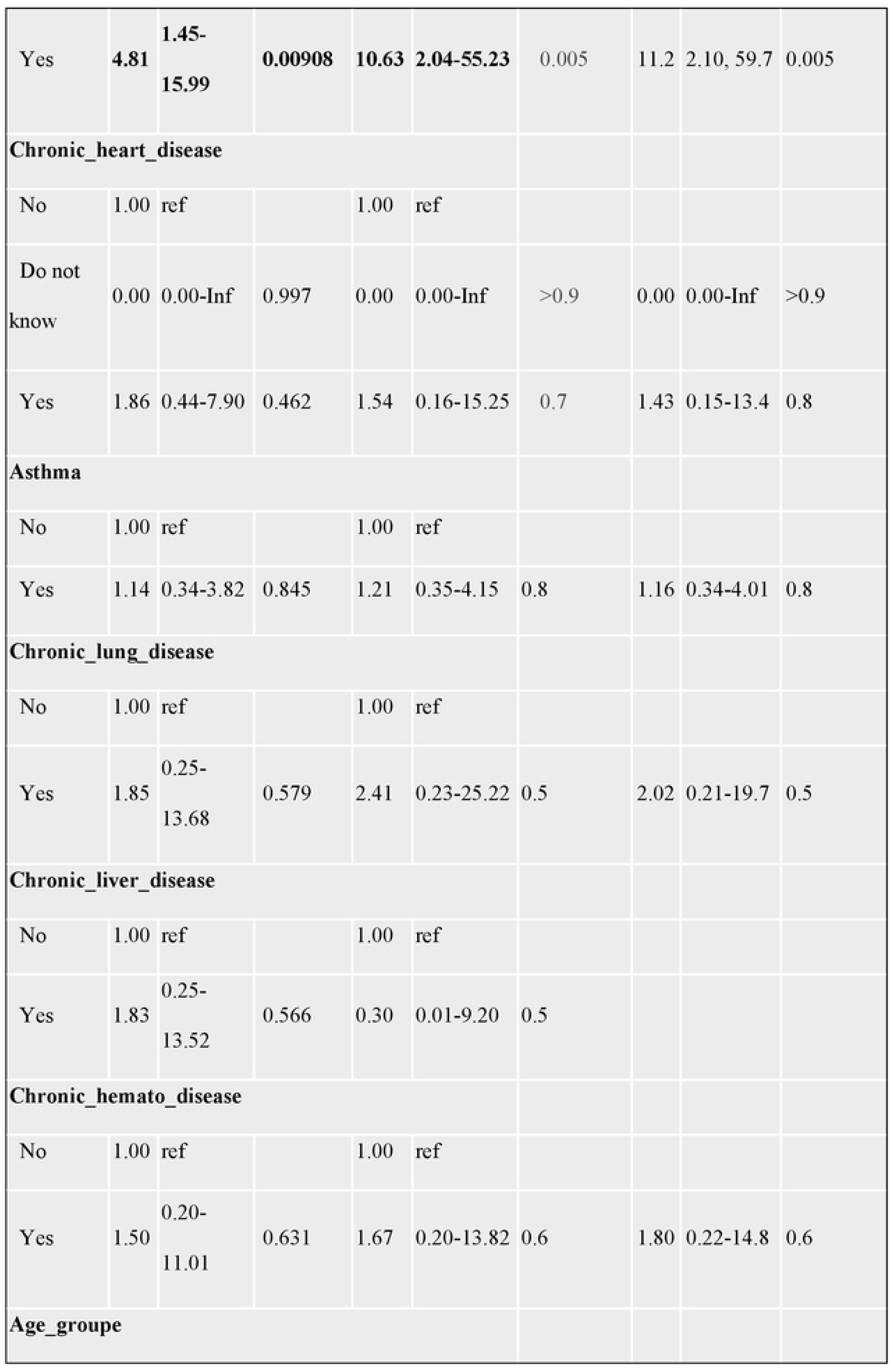
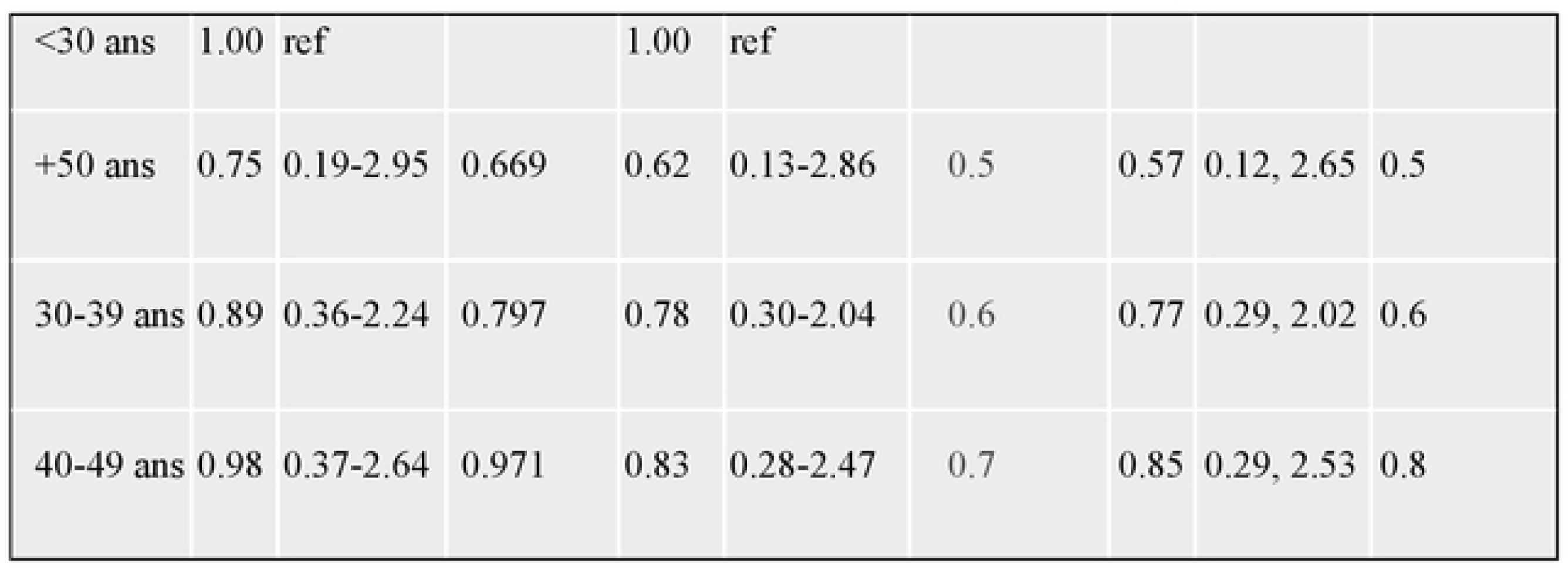
Risk factors associated with reporting covid symptoms among health workers in Niger after contact with a covid-19 patient.

For clinical disease reported during the first 45 days after contact (Table VI), the models highlighted diabetes as the only factor associated with Covid symptom (models I, II, III).

**Table VI :**
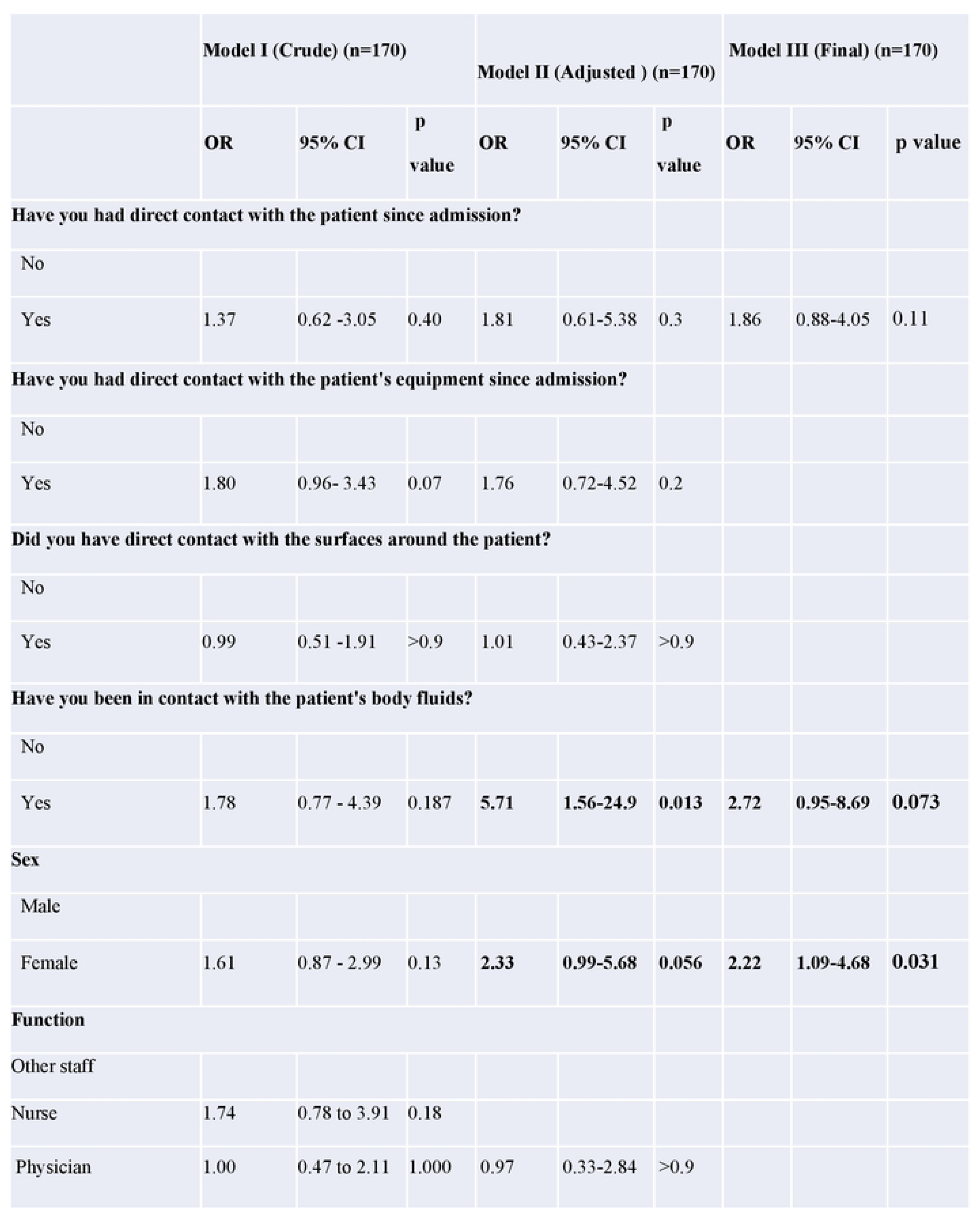
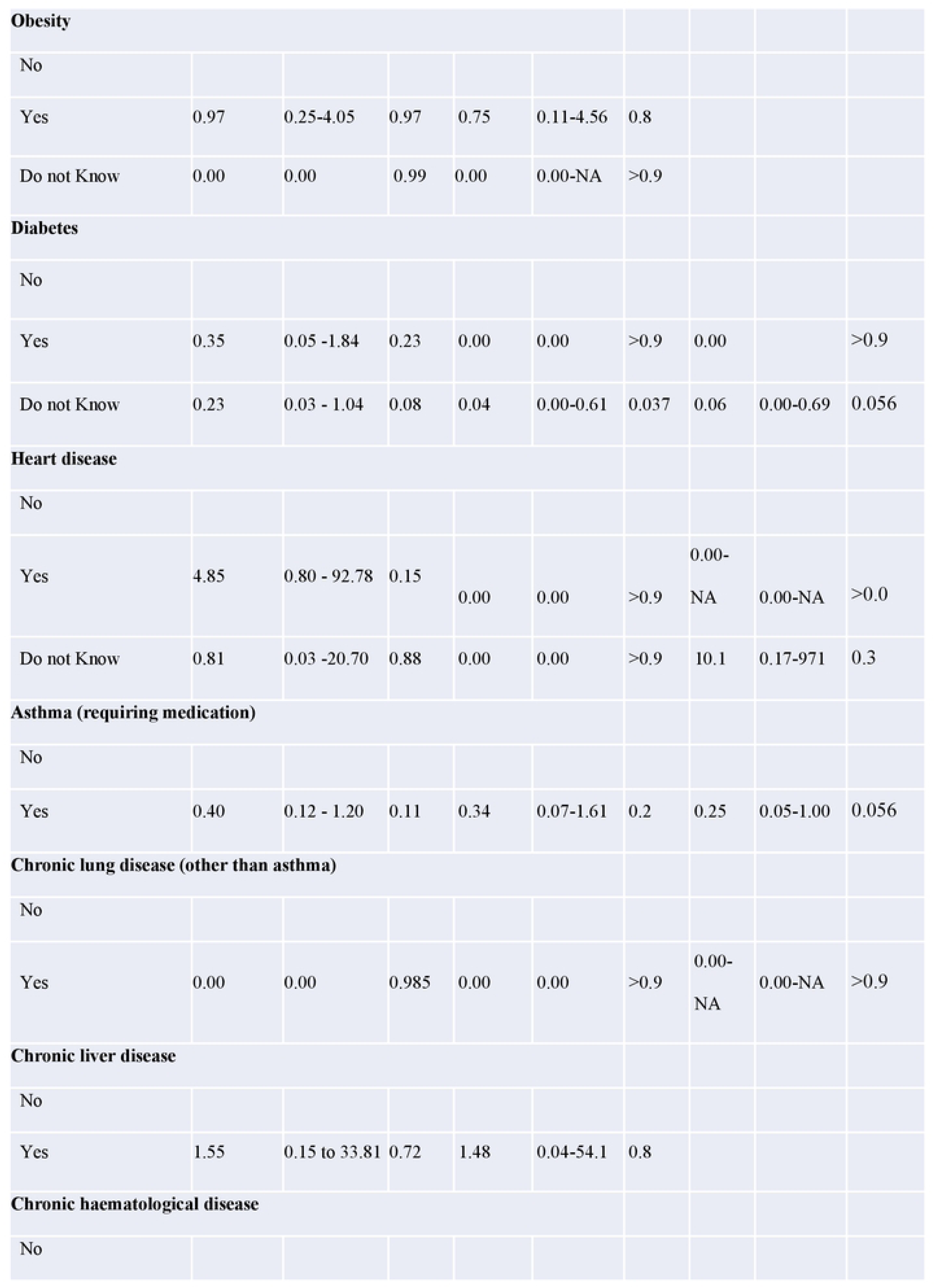
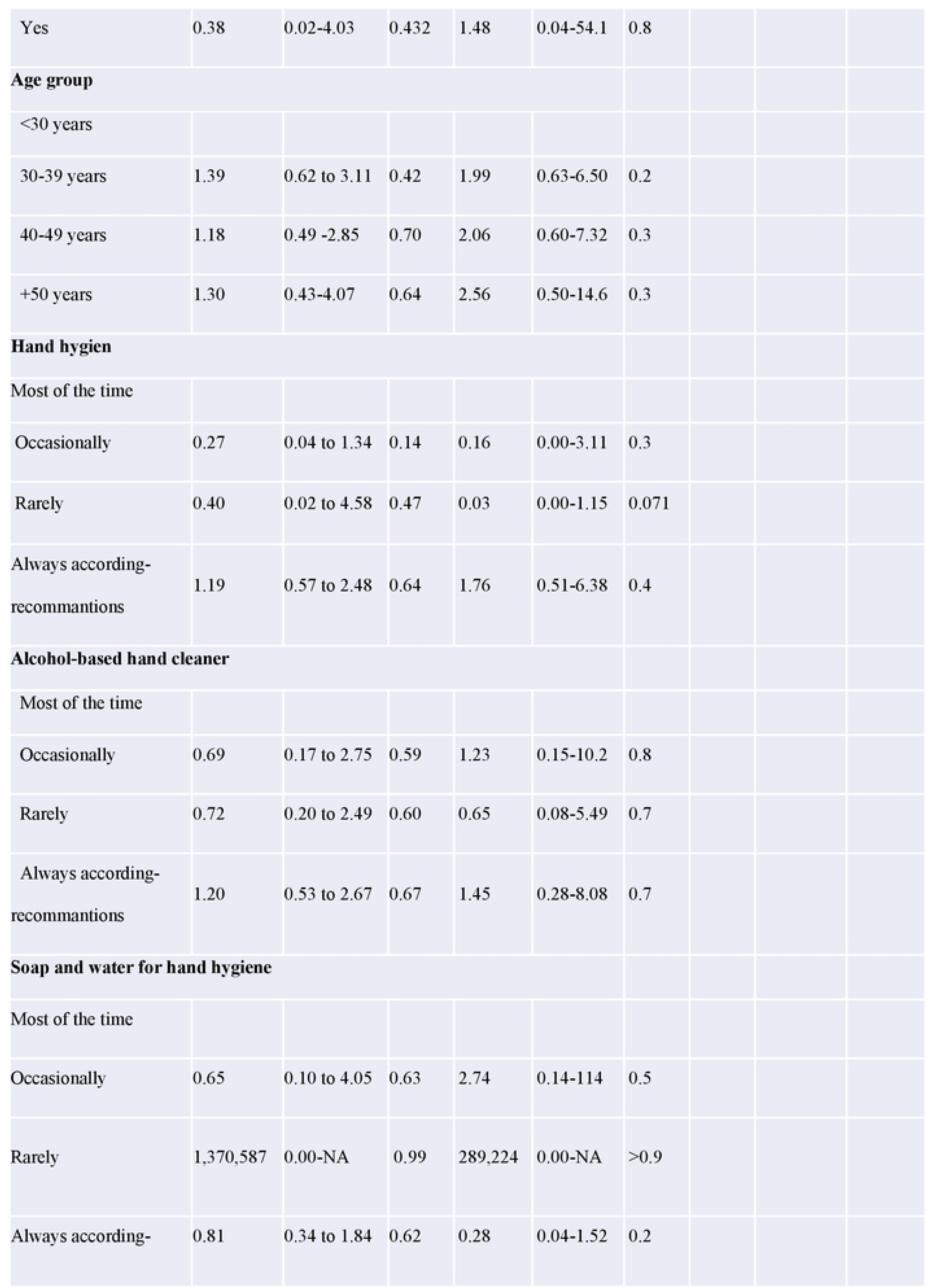
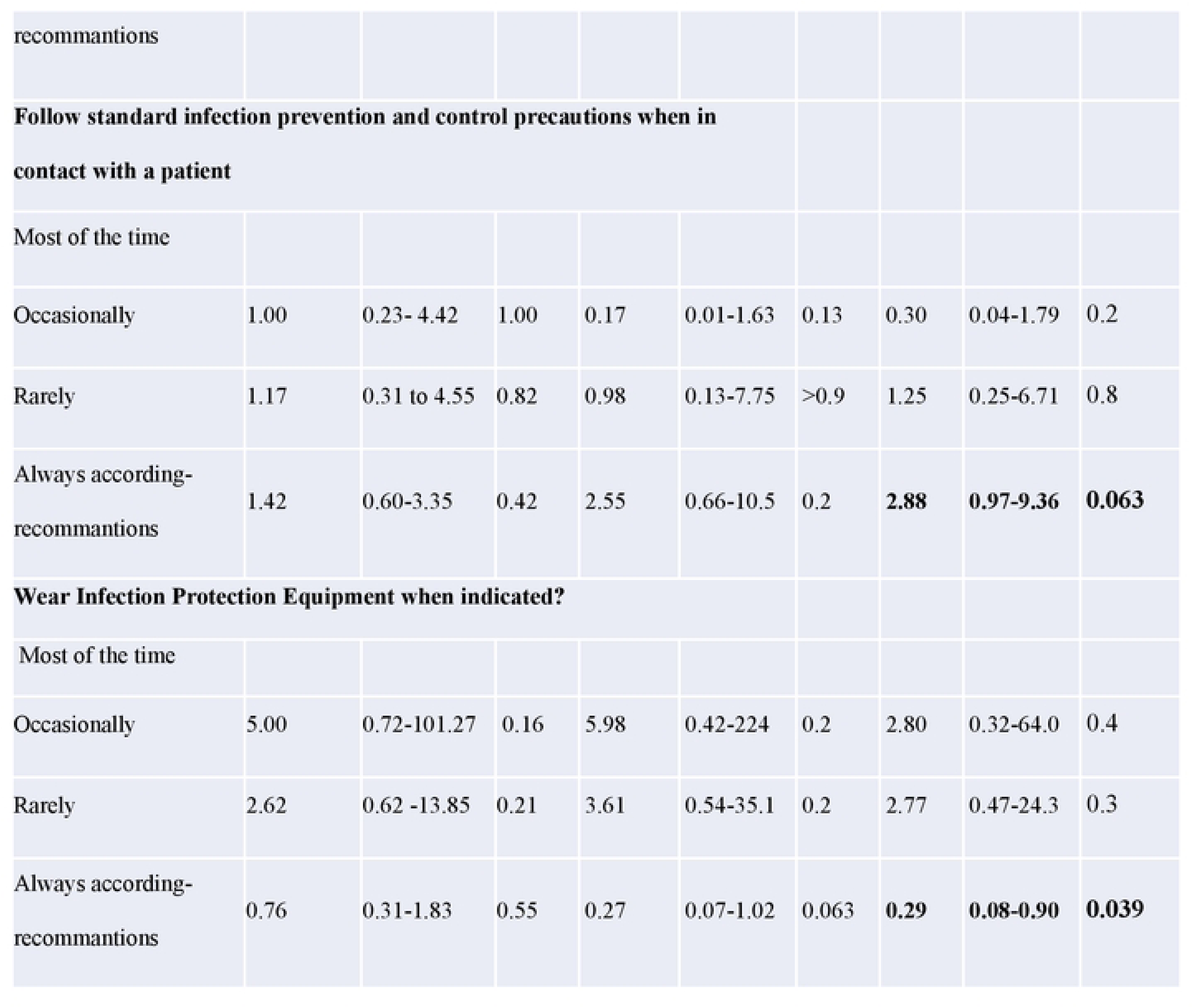
Overall risk factors associated with COVID-positive status among health workers in Niger.

## 4. Discussion

This prospective cohort study aims to analyze risk factor for COVID-19 among health workers during the first wave of the SARS COVID-2 epidemic in Niger. It focused on workers who were in contact with the first confirmed cases of SARS CoV-2 in Niamey. The strategy was to follow them for 30 days after contact. This research was conducted in Niamey, which was the epicenter of the epidemic in Niger. During this time, and despite awareness resulting for the previous Ebola epidemic in the West African region, good clinical practices were not well established in dispensaries and first line heath structures. In this context, in case of positive patient attending the ward, all the workers from medical doctors to technicians in laboratories and cleaners can be impacted. The main questions to address were which type of activity was at major risk, and in the other hand, which practices can be protective.

The biological methods used were polymerase chain reaction, which is the gold standard for diagnosis of infection, and ELISA for serology to demonstrate exposure to SARS CoV-2. However, as it was not possible to practice regular PCR test to all the workers at fixed date, this tests was only performed when clinical signs were declared.

In this study covering one month of follow-up, the percent of people becoming PCR positive was 28.8% with a very high difference between gender, i.e. 64.7% among females and 5.29% among males. Our result is superior to that a similar study performed in Egypt between April and May 2020 on 4040 healthcare workers where 170 (4.2%; 95%) were infected with SARS-CoV-2 (II) (11). In a study conducted from March 2020 to December 2020 at Med Tlatli Hospital in Nabeul, Tunisia, the rate of SARS-CoV-2 positivity among health care personnel was 14.4%. The frequency of infection was 15.4% (12). En Algérie, une étude transversale descriptive menée du 01/03/2020 au 31/08/2021 en Algérie avait enregistré une prévalence de 15 % de l’infection COVID-19 (13).

SARS CoV-2 seroprevalence reaches 55.9% after one month, which is much higher than in Nigeria 45.10% (14), Zimbabwe 8.9% (15), UK 24.4% (16), and The Netherlands 21.1% (17). This high rate of contamination of health workers during the first wave of COVID epidemic, underlines the poor level of knowledge that they have about the disease. and also, their poor access to individual protection.

Due to the low age of the working population in Niger, most of the workers didn’t harbored comorbidity increasing their risk of disease. However, 21.2% of the health workers reported one comorbidity and 6.56% several. The most recorded risk factors were asthma (7.33%), obesity (5.01%), heart disease (3.80%) followed by chronic lung disease (2.31%). Chronic liver disease was significantly (p= 0.034) associated with positive COVID rRT-PCR as already reported.

In the univariate analysis, we noticed a significant difference in the proportion of positive PCRs according to function (p=0.024), and structure to which the health worker belonged. According to literature, it is clear that the risk varies by occupation and workstation. However, awareness could be more important than equipment. Indeed, a study conducted in the United Kingdom reported a higher seroprevalence among housekeepers (34.5%) and those working in acute medicine (33%) or general internal medicine (30.3%) than in intensive care units welcoming most of the patients (14.8%) (15). Similar results were reported from Egypt where 39.6% of positive were nurses, 39.0% were physicians but 21.4% were non-medical staff (11). Our results are consistent with another study conducted in Niger where health workers accounted for 12.8% of confirmed cases, followed by students (9.8%) (18), with a higher risk of death among general practitioners and mental health nurses (19). Frontline health care workers had at least a threefold increased risk of positive COVID-19 test and infection, compare with the general community (20). Awareness and knowledge of the disease, seem to be the main pitfalls in protection of health workers. In the same line, hand washing (p=0.019), and alcohol-base hand hygiene (p<0.001) were also mainly associated with negative serology, in the opposite of contact with contaminated material (p=0.069). These results are consistent with those of Li Ran *et al*. in Wuhan, China demonstrating that contamination of health care workers is generally due to hand contamination after contact with either patients or fomites (21). These results highlight the importance of hand hygiene and alcohol-base hand hygiene. In dispensaries, personal protective equipment are usually limited to gloves when present. They can offer an effective protection and according to Ng-Kamstra *et al*. more complex PPE protocols do not necessarily offer better one. They sometime may even increase the risk of self-contamination when removing the them (22).

During infectious diseases outbreaks, implementation of prevention and control (IPC) is of great importance in health care facilities especially with regard to the protection of health care workers . In Niger, and more specifically in Niamey, which was the epicenter of the epidemic, handwashing and water devices were installed in all health facilities. In this study the average score of use ranged from 52% to 54% and 55% in tertiary health facilities, integrated health centers and private health facilities respectively. However, the WHO recommended technique of hand washing or hand rubbing with hydro alcoholic gel were not sufficiently mastered by the workers (6).

More commitment and sensitization of health workers in the front line in the fight against pathology is needed. As we all know, hand washing not only significantly reduces residual viruses or bacteria on the hands but also reduces the sources of contamination.

## 5. Conclusion

The identification of risk factors in the health care setting is essential and allows policy makers to adopt appropriate strategies against this disease. Preventive measures such as hygiene in general must be privileged in our constraints in addition to personal protective equipment.

**Conflicts of interest:** the authors declare no conflicts of interest.

**Funding:** This study was funded by the French Ministry of Europe and Foreign Affairs via the “MediLabSecure” project coordinated by Pasteur International Network Association.

## Data Availability

The data cannot be shared due to its confidentiality. They are available at our institution, the Center for Medical and Health Research (CERMES) (contact via BP: 10887, Tel +22720752040/45).

## Acknowledgements

**Acknowledgements:** To the Ministry of Public Health, Population and Action, and to the health care facilities for accepting the implementation of this study in their respective centers. To MediLabSecure for funding the study reported in this article. To all CERMES staff who in one way or another participated in the realization of this study.

**Contribution to the study:** HLR, RJ participated in the conception and design of the study, RHL, RJ, MM, FAA participated in the data analysis, ASK, LA, AFC, IK participated in the analysis of the samples in the laboratory, FAA, II, ZA, BAS, IG, AS, ZAA, AM collected field data, RH, RJ, MM drafted the manuscript, GAMB, IG, SM analysis of the rough draft, HLR, JR, MM, IML, AL critically reviewed the manuscript.

All authors read and approved the final version of this manuscript.

## Références bibliographiques

1. Pneumonia of unknown cause – China [Internet]. [cité 16 août 2022]. Disponible sur: https://www.who.int/emergencies/disease-outbreak-news/item/2020-DON229

2. Kantorski LP, de Oliveira MM, Alves PF, Treichel CA dos S, Wünsch CG, dos Santos LH, et al. Intention to leave Nursing during the COVID-19 pandemic. Rev Lat Am Enfermagem. 30:e3613.

3. Biancolella M, Colona VL, Mehrian-Shai R, Watt JL, Luzzatto L, Novelli G, et al. COVID-19 2022 update: transition of the pandemic to the endemic phase. Hum Genomics. 1 juin 2022;16:19.

4. Sougou NM, Diouf JB, Diallo AA, Seck I. Analyse des risques perçus des prestataires de santé en milieu hospitalier dans le cadre de la pandémie à COVID-19: une étude qualitative dans le Centre Hospitalier Roi Baudoin de Guédiawaye, lors de la réception du 1er cas communautaire du Sénégal. Pan Afr Med J. 13 oct 2020;37(Suppl 1):23.

5. Rosenthal PJ, Breman JG, Djimde AA, John CC, Kamya MR, Leke RGF, et al. COVID-19: Shining the Light on Africa. Am J Trop Med Hyg. juin 2020;102(6):1145–8.

6. Baissa AM, Hamani S, Ali M, Mouako AL, Anya BPM, Wiysonge CS. La lutte contre le COVID-19 au Niger: ĺévaluation de la prévention et contrôle des infections dans les formations sanitaires de la communauté urbaine de Niamey. Pan Afr Med J. 11 nov 2020;37(Suppl 1):35.

7. Consolider la riposte au Niger, deux ans après le premier cas de COVID-19 dans le pays [Internet]. OMS | Bureau régional pour l’Afrique. 2022 [cité 15 févr 2023]. Disponible sur: https://www.afro.who.int/fr/countries/niger/news/consolider-la-riposte-au-niger-deux-ans-apres-le-premier-cas-de-covid-19-dans-le-pays

8. Gómez-Ochoa SA, Franco OH, Rojas LZ, Raguindin PF, Roa-Díaz ZM, Wyssmann BM, et al. COVID-19 in Health-Care Workers: A Living Systematic Review and Meta-Analysis of Prevalence, Risk Factors, Clinical Characteristics, and Outcomes. Am J Epidemiol. 4 janv 2021;190(1):161–75.

9. WHO-2019-nCoV-health_workforce-2020.1-fre.pdf [Internet]. [cité 17 août 2022]. Disponible sur: https://apps.who.int/iris/bitstream/handle/10665/341555/WHO-2019-nCoV-health_workforce-2020.1-fre.pdf

10. Tabah A, Ramanan M, Laupland KB, Buetti N, Cortegiani A, Mellinghoff J, et al. Personal protective equipment and intensive care unit healthcare worker safety in the COVID-19 era (PPE-SAFE): An international survey. J Crit Care. oct 2020;59:70–5.

11. Mostafa A, Kandil S, El-Sayed MH, Girgis S, Hafez H, Yosef M, et al. Universal COVID-19 screening of 4040 health care workers in a resource-limited setting: an Egyptian pilot model in a university with 12 public hospitals and medical centers. Int J Epidemiol. 23 oct 2020;dyaa173.

12. Mrazguia C, Aloui H, Fenina E, Boujnah A, Azzez S, Hammami A. Ĺinfection par le COVID-19 chez le personnel de santé à ĺHôpital Régional de Nabeul : épidémiologie et circonstances de transmission. PAMJ - One Health [Internet]. 4 mars 2021 [cité 15 févr 2023];4(11). Disponible sur: https://www.one-health.panafrican-med-journal.com/content/article/4/11/full

13. Ghomari O, Merad S, Moulessehoul F, Merad Y. Infection COVID-19 chez le personnel de santé : expérience du service de médecine du travail d’un CHU de l’ouest Algérien. Arch Mal Prof Environ. août 2022;83(4):368.

14. Olayanju O, Bamidele O, Edem F, Eseile B, Amoo A, Nwaokenye J, et al. SARS-CoV-2 Seropositivity in Asymptomatic Frontline Health Workers in Ibadan, Nigeria. Am J Trop Med Hyg. janv 2021;104(1):91–4.

15. Rusakaniko S, Sibanda EN, Mduluza T, Tagwireyi P, Dhlamini Z, Ndhlovu CE, et al. SARS-CoV-2 Serological testing in frontline health workers in Zimbabwe. PLoS Negl Trop Dis. mars 2021;15(3):e0009254.

16. Shields A, Faustini SE, Perez-Toledo M, Jossi S, Aldera E, Allen JD, et al. Original research: SARS-CoV-2 seroprevalence and asymptomatic viral carriage in healthcare workers: a cross-sectional study. Thorax. déc 2020;75(12):1089.

17. Bouwman M, van Osch F, Crijns F, Trienekens T, Mehagnoul-Schipper J, van den Bergh JP, et al. SARS-CoV-2 seroprevalence in healthcare workers of a teaching hospital in a highly endemic region in the Netherlands after the first wave: a cross-sectional study. BMJ Open. 18 oct 2021;11(10):e051573.

18. Tchole AIM, Li ZW, Wei JT, Ye RZ, Wang WJ, Du WY, et al. Epidemic and control of COVID-19 in Niger: quantitative analyses in a least developed country. J Glob Health. 10(2):020513.

19. Bandyopadhyay S, Baticulon RE, Kadhum M, Alser M, Ojuka DK, Badereddin Y, et al. Infection and mortality of healthcare workers worldwide from COVID-19: a systematic review. BMJ Glob Health. 4 déc 2020;5(12):e003097.

20. Nguyen LH, Drew DA, Graham MS, Joshi AD, Guo CG, Ma W, et al. Risk of COVID-19 among front-line health-care workers and the general community: a prospective cohort study. Lancet Public Health. sept 2020;5(9):e475–83.

21. Ran L, Chen X, Wang Y, Wu W, Zhang L, Tan X. Risk Factors of Healthcare Workers with Corona Virus Disease 2019: A Retrospective Cohort Study in a Designated Hospital of Wuhan in China. Clin Infect Dis Off Publ Infect Dis Soc Am. 17 mars 2020;ciaa287.

22. Ng-Kamstra J, Stelfox HT, Fiest K, Conly J, Leigh JP. Perspectives sur l’équipement de protection individuelle dans les établissements de soins actifs durant la pandémie de COVID-19. CMAJ Can Med Assoc J. 7 déc 2020;192(49):E1785–90.

